# Sleep Duration and Long-Term Mortality After Stroke: A Nationwide Analysis

**DOI:** 10.1101/2023.12.11.23299835

**Authors:** Sara Hassani, Bruce Ovbiagele, Daniela Markovic, Amytis Towfighi

## Abstract

**Background:** Sleep duration is a key marker of ideal cardiovascular health in the American Heart Association’s new Life’s Essential 8 construct, but studies on outcomes in stroke survivors are scarce. We assessed the association between sleep duration and mortality among self-reported stroke survivors in a nationally representative U.S. sample.

**Methods:** Cross-sectional data (2005–2018) from the National Health and Nutrition Examination Surveys database in patients aged ≥18 (n=42,143) with a self-reported history of stroke (n=1,488) were linked to the 2019 National Death Index to determine the association between nightly sleep duration and mortality. Relationships between sleep duration (short: <7 hours, normal: 7-8 hours, long: >8 hours) and demographic characteristics were assessed. Relationships between sleep duration and mortality (all-cause and cardiovascular causes) were evaluated adjusting for demographic and clinical variables, with multivariable Cox regressions.

**Results:** Among stroke survivors, prevalence of short sleep duration increased with younger age while prevalence of long sleep duration increased with older age (p<0.001). There were no significant sex differences in sleep duration. Long sleep duration was associated with higher all-cause mortality in the unadjusted model (HR 1.82, 1.44-2.31, p<0.0001). After adjustment for co-variates, (HR 1.36, 1.08-1.71, p=0.01), the association attenuated, but remained significant (HR 1.30, 1.02-1.65, p=0.03). Sensitivity analysis further verified the reliability of this conclusion. Short sleep duration was not associated with all-cause or cardiovascular mortality after adjusted analyses. There were no significant associations between long sleep duration and cardiovascular mortality.

**Conclusion:** Long sleep duration is independently associated with higher risk, all-cause mortality after stroke.

## Introduction

Sleep duration is now considered an essential component for ideal brain and heart health according to the American Heart Association’s Life’s Essential 8 construct^1^, emphasizing the need to understand the implications of abnormal sleep patterns. In prospective cohort studies, among myocardial infarction survivors or those with an established history of coronary artery disease, both short (< 8 hours)^2–4^ and long sleep duration (>8 hours)^2,3^ have been shown to be independent and strong predictors of all-cause mortality. Shortened^5–9^ or prolonged^5–7,9–11^ sleep length in the general population has been associated with increased risk of stroke incidence.^9^ Among survivors of stroke, sleep duration has also been strongly associated with poor functional status and neurocognitive impairment.^12,13^ But whether the duration of sleep after a stroke also affects mortality outcomes remains unknown.

To address this knowledge gap, this study aimed to assess whether sleep duration is associated with an independent, higher risk of all-cause or cardiovascular mortality among stroke survivors in a large nationally representative United States sample. Characterizing relationships between post-stroke sleep and mortality may help guide targeted intervention and policy development to improve health outcomes at the patient and population levels.

## Methods

### Study Population

The National Health and Nutrition Examination Surveys (NHANES) gathers data using two-year cycles on the nutritional and health status of the US people and is conducted by the National Center for Health Statistics, a branch of the Centers for Disease Control and Prevention. Individuals participating in NHANES are interviewed in their homes and examined at mobile examination centers. Signed informed consent is obtained from each person during the home interview. NHANES uses a complex, multistage probability sampling design to select a sample representative of the civilian noninstitutionalized household population of the United States.^14^ At different time points, subgroups of particular public health interest are oversampled to increase the reliability and precision of estimates of health status indicators for those population subgroups. Weighting schemes allow estimates from those subgroups to be combined to obtain national estimates reflective of the relative proportions of those groups in the overall population.^14^ There are approximately 10,000 individuals in each cycle of this nationwide, cross-sectional survey.^15^

NHANES data sets were downloaded from the website of the National Center for Health Statistics (http://www.cdc.gov/nchs) for the survey years 2005 – 2018. Our study used the NHANES data for the 2005–2006, 2007–2008, 2009–2010, 2011–2012, 2013–2014, 2015-2016, and 2017-2018 cycles. The National Center for Health Statistics Research Ethics Review Board approved the 2005–2018 NHANES protocols.

The study excluded individuals younger than 18 years, or those with missing values in the sleep duration variable required for the analyses.

### Measures

#### Stroke

History of stroke was diagnosed by a positive answer to the question, “Has a doctor or other health professional ever told you that you had a stroke?”

#### Sleep Duration

During the five cycles of NHANES from 2005 – 2018, sleep duration was assessed by the question “How much sleep do you usually get at night on weekdays or workdays?” Participants were asked to indicate how many hours per day they usually sleep. Sleep duration was divided for the analyses into 3 commonly cited categories: short sleep, <7 hours per day; normal sleep, 7 to 8 hours of sleep; and long sleep, ≥8 hours of sleep.^16,17^

For a sensitivity analysis, we repeated the main analysis with the reference categories^18^ of sleep duration being defined instead as: short (less than 6 hours), normal (6 to 9 hours), and long (over 9 hours), respectively.

#### Mortality

The data of serial NHANES 2005 – 2018 cycles was linked to the National Death Index through December 31, 2018. The primary study outcome variable was all-cause mortality, analyzed as a time to event outcome (event was deceased from all-causes versus alive). The secondary outcome variable was cardiovascular mortality, analyzed as a time to event outcome (event was deceased attributable to cardiovascular causes versus alive or deceased attributable to competing non-cardiovascular causes). Death from cardiovascular disease included deaths from any heart disease, cerebrovascular cause, atherosclerosis, or hypertension (UCOD-113 codes 054 to 074). Stroke mortality (deaths from any cerebrovascular cause, UCOD-113 code 070) was not used as a primary outcome as it was too rare to formally control for covariates.

#### Covariates

Information on potential confounders, including education, weight, body mass index (BMI), and history of hypertension, myocardial infarction, cancer, diabetes mellitus, and depression were obtained from the questionnaires completed by the participants, and considered binary variables. Demographic characteristics were also collected: sex (male, female); race (Mexican American, other Hispanic, white non-Hispanic, black non-Hispanic, and other); education level (less than high school, high school graduate or some college, and college graduate or above); and family poverty income ratio (PIR, a ratio of family income to poverty threshold of less than or equal to 200%, greater than 200%, or unknown). Age was stratified into three groups: 18-44, 45-64, and 65 or greater years of age.

Hypertension was defined by self-reported physician diagnosis, self-reported current antihypertensive medication use, or a mean of the first 3 blood pressure readings >140 mm Hg systolic or 90 mm Hg diastolic. Diabetes mellitus was defined by self-reported physician diagnosis, self-reported current medical therapy, or glycosylated hemoglobin >7%. History of myocardial infarction (MI) was defined by self-reported physician diagnosis of “heart attack.” History of cancer was defined by self-reported physician diagnosis of “cancer.” Depression was determined by a Patient Health Questionnaire-9 score ≥10.

### Statistical Analysis

Prevalence of sleep duration was estimated and compared across age, sex, and race/ethnic groups using the Rao-Scott Chi-square test. Prevalence estimates were weighted using the NHANES interview sample weight to adjust for differential probabilities of selection and nonresponse. Survey adjusted Cox proportional hazard regression models were used to analyze the association of short and long sleep duration on all-cause mortality. For assessments of sleep duration on cardiovascular mortality, the cause specific competing risk model was used (implemented via “proc surveyphreg” in SAS). We performed a series of nested models to assess the influence of specific covariates on the above-mentioned associations. Model 1 was the unadjusted model; Model 2 was adjusted for demographic variables (age, sex, race/ethnicity, education level, and family PIR); Model 3 was adjusted for the above demographic and additional clinical variables (BMI, history of coronary artery disease, myocardial infarction, cancer, hypertension, diabetes, and PHQ-9 score of ≥10 consistent with depression). All models allowed for interactions of sleep duration with each of the covariates by including the appropriate interaction terms to the models. Data analyses were conducted using SAS version 9.4 (SAS Institute Inc, Cary, North Carolina, USA). Statistical hypotheses were tested using *p* < 0.05 as the level of statistical significance.

## Results

During the five cycles of NHANES from 2005 – 2018, there were a total of 42,143 participants aged >= 18 years included. History of stroke was identified by self-report in 1,507 out of 42,143 participants. An additional 19 participants were excluded because of missing values in the sleep duration variable required for the analyses. A total of 1,488 participants were included in the analyses (**Figure 1**).

Characteristics of the 1,488 stroke survivors included in the study are summarized in **Table 1**. Among stroke survivors, prevalence of short sleep duration increased with younger age while prevalence of long sleep duration increased with older age (p<0.001). There were no significant differences in sleep duration between men and women (p=0.18). Prevalence of short sleep duration was higher among Hispanic and Non-Hispanic Black individuals compared to non-Hispanic White individuals (p<0.0001). Stroke survivors with long sleep duration (vs. normal) were more likely to be older and overweight, and those with short sleep duration (vs. normal) were more likely to be younger, non-White, and obese (age p<0.001, race/ethnicity p<0.001, BMI p=0.001).

**Table 1.**
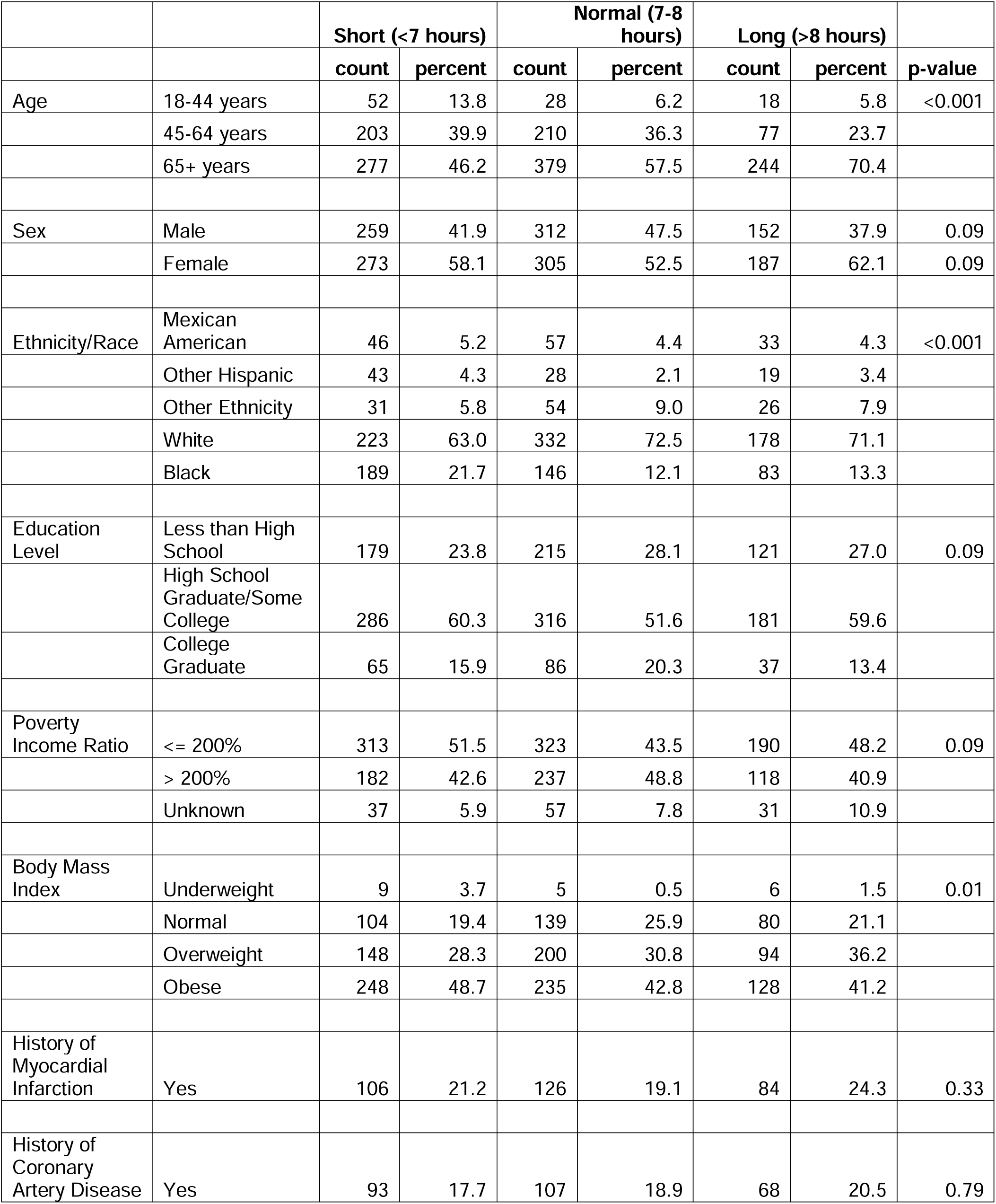
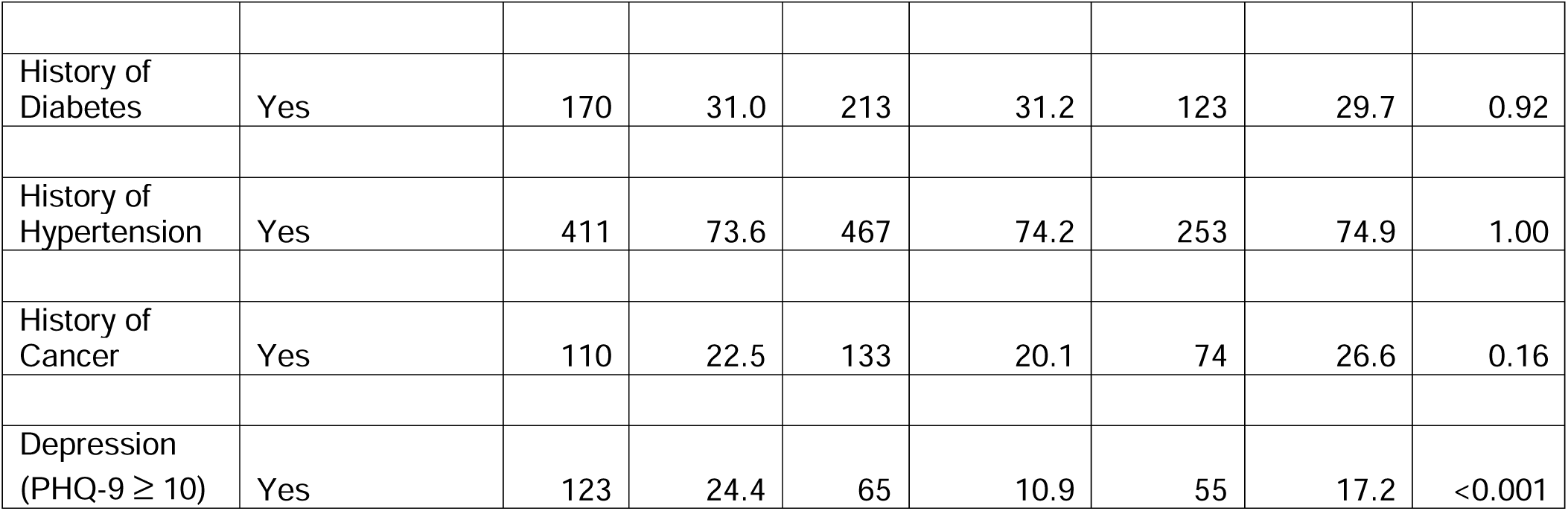
Demographic Features of Sleep Duration Categories, NHANES 2005-2018.

Long sleep duration was associated with higher all-cause mortality in the unadjusted model (HR 1.82, 1.44-2.31, p<0.0001). After adjusting for both demographic variables (HR 1.36, 1.08-1.71, p=0.01) and comorbidities, the association attenuated, but remained significant (HR 1.30, 1.02-1.65, p=0.03). Sensitivity analysis (with long sleep defined as >9 hours) further verified the reliability of this conclusion (HR 1.42, 1.10-1.90, p=0.02) in the adjusted models **(Table 4).** Survival curves are shown in **Figures 2 and 3**.

There were no significant associations between short sleep duration and all-cause or cardiovascular mortality (**Tables 2 and 3)**. The sensitivity analyses, with alternate reference categories of sleep duration being defined as: short – less than 6 hours, normal – 6 to 9 hours, and long – over 9 hours,^18^ also did not reveal any statistically significant associations between short sleep and all-cause mortality **(Table 4).** Short sleep duration on sensitivity analysis was associated with lower cardiovascular mortality in the unadjusted model (HR 0.51, 0.32-0.82, p=0.01), but the effect was no longer significant after adjusting for demographic variables (p=0.50) (**Table 5).** There were no significant associations between long sleep duration and cardiovascular mortality.

**Table 2.** All-Cause Mortality after Stroke Stratified by Sleep Duration.

|  |  |  |  |
| --- | --- | --- | --- |
| **Model 1: Unadjusted** |  |  |  |
| **Sleep Duration** | **Hazard Ratio** | **(95% CI)** | **P-value** |
| Long (> 8 hours) | 1.82 | (1.44-2.31) | <0.0001 |
| Short (< 7 hours) | 0.79 | (0.61-1.02) | 0.08 |
| Normal (7-8 hours) | 1.00 |  |  |
| **Model 2: Adjusted for Demographic Variables\*** |  |  |  |
| Long (> 8 hours) | 1.35 | (1.08-1.71) | 0.01 |
| Short (< 7 hours) | 0.98 | (0.79-1.21) | 0.84 |
| Normal (7-8 hours) | 1.00 |  |  |
| **Model 3: Adjusted for Demographic and Clinical Variables\*\*** |  |  |  |
| Long (> 8 hours) | 1.30 | (1.02-1.65) | 0.03 |
| Short (< 7 hours) | 0.97 | (0.78-1.22) | 0.82 |
| Normal (7-8 hours) | 1.00 |  |  |

\*Adjusted for age, sex, and race
\*\* Adjusted for age, sex, race, highest education level, body mass index, and history of stroke, myocardial infarction, hypertension, diabetes, coronary artery disease, cancer, and depression

**Table 3.** Cardiovascular Mortality after Stroke Stratified by Sleep Duration.

|  |  |  |  |
| --- | --- | --- | --- |
| **Model 1: Unadjusted** |  |  |  |
| **Sleep Duration** | **Hazard Ratio** | **(95% CI)** | **P-value** |
| Long (> 8 hours) | 1.35 | (0.86-2.99) | 0.14 |
| Short (< 7 hours) | 0.73 | (0.30-1.35) | 0.23 |
| Normal (7-8 hours) | 1.00 |  |  |
| **Model 2: Adjusted for Demographic Variables\*** |  |  |  |
| Long (> 8 hours) | 0.98 | (0.57-2.15) | 0.76 |
| Short (< 7 hours) | 0.92 | (0.35-1.71) | 0.53 |
| Normal (7-8 hours) | 1.00 |  |  |
| **Model 3: Adjusted for Demographic and Clinical Variables\*\*** |  |  |  |
| Long (> 8 hours) | 0.94 | (0.63-1.40) | 0.75 |
| Short (< 7 hours) | 0.91 | (0.61-1.34) | 0.61 |
| Normal (7-8 hours) | 1.00 |  |  |

\*Adjusted for age, sex, and race
\*\* Adjusted for age, sex, race, highest education level, body mass index, and history of stroke, myocardial infarction, hypertension, diabetes, coronary artery disease, cancer, and depression

**Table 4.**
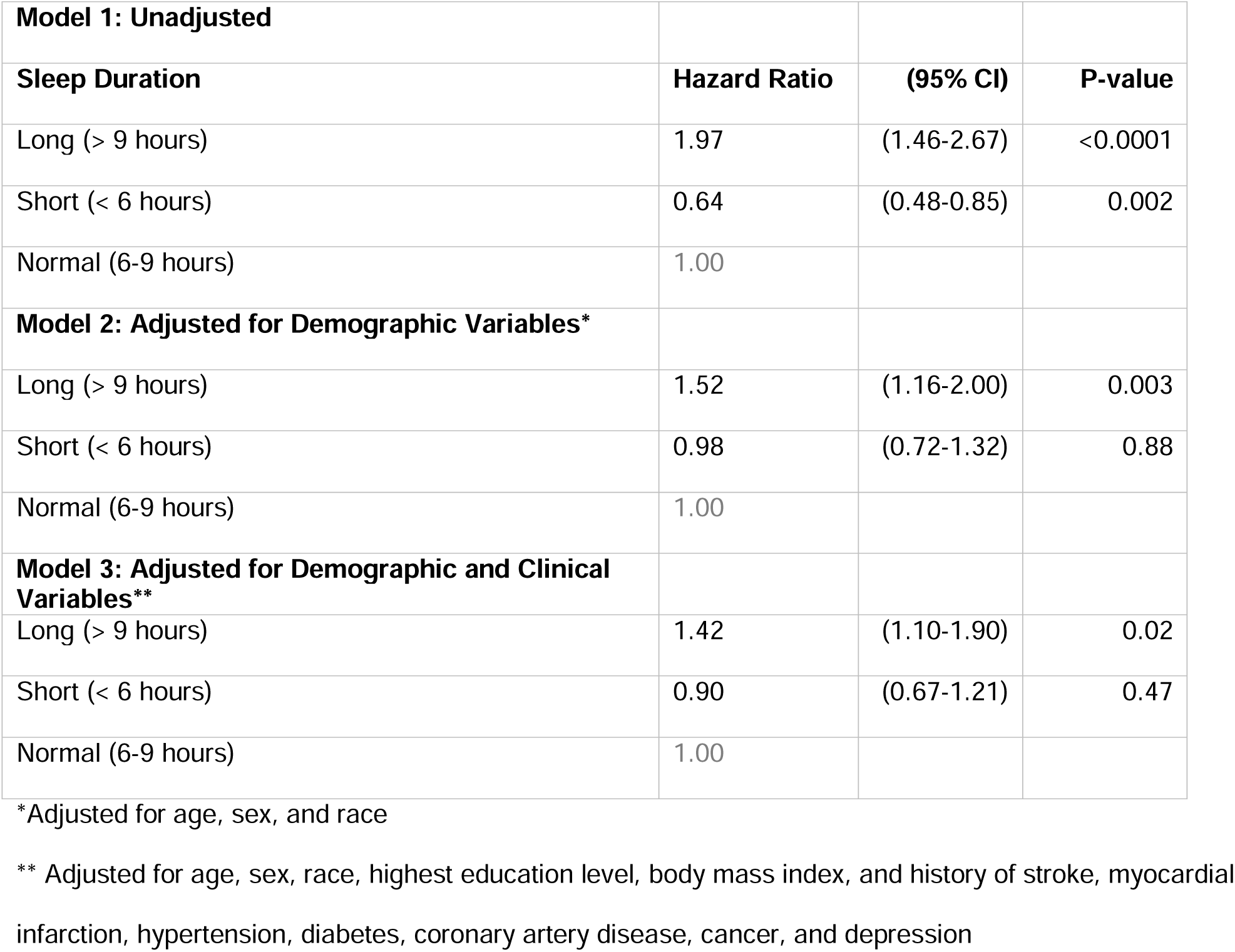
Hazard Ratios of All-Cause Mortality According to Sleep Duration Among Stroke Survivors, Sensitivity Analysis.

**Table 5.** Hazard Ratios of Cardiovascular Mortality According to Sleep Duration Among Stroke Survivors, Sensitivity Analysis.

|  |  |  |  |
| --- | --- | --- | --- |
| **Model 1: Unadjusted** |  |  |  |
| **Sleep Duration** | **Hazard Ratio** | **(95% CI)** | **P-value** |
| Long (> 9 hours) | 1.55 | (0.86-2.78) | 0.15 |
| Short (< 6 hours) | 0.51 | (0.32-0.82) | 0.01 |
| Normal (6-9 hours) | 1.00 |  |  |
| **Model 2: Adjusted for Demographic Variables\*\*** |  |  |  |
| Long (> 9 hours) | 1.12 | (0.66-1.92) | 0.76 |
| Short (< 6 hours) | 0.84 | (0.52-1.38) | 0.50 |
| Normal (6-9 hours) | 1.00 |  |  |
| **Model 3: Adjusted for Demographic and Clinical Variables\*\*** |  |  |  |
| Long (> 9 hours) | 1.42 | (0.61-1.82) | 0.85 |
| Short (< 6 hours) | 0.90 | (0.47-1.28) | 0.32 |
| Normal (6-9 hours) | 1.00 |  |  |

\*Adjusted for age, sex, and race
\*\* Adjusted for age, sex, race, highest education level, body mass index, and history of stroke, myocardial infarction, hypertension, diabetes, coronary artery disease, cancer, and depression

## Discussion

The primary finding of this study, the largest and most up-to-date survey on sleep duration representative of a national sample of non-institutionalized stroke survivors in the United States, is an independent, higher risk association between long sleep duration and all-cause mortality after stroke. No associations between long sleep duration and cardiovascular mortality or short sleep with all-cause or cardiovascular mortality were noted.

Many epidemiologic studies and meta-analyses in the general population have shown that both short and long reported sleep durations are independently associated with all-cause and cardiovascular mortality^7,19–21^ as well as cardiovascular events,^7,22^ though these U-shaped associations have not been consistent.^23,24^ Prior studies on sleep duration and mortality in a population of known MI survivors have demonstrated that both short and long durations of sleep are independent predictors of all-cause mortality,^2,3^ and some also have shown a similar pattern for cardiovascular mortality, though only for short, and not long sleep duration.^3^ However, this is the first study to our knowledge to investigate the impact of sleep duration on outcomes in a population with prior stroke.

In light of studies of myocardial infarction survivors that have reported a U-shaped rather than a J-shaped association with all-cause mortality,^3^ the discrepancy of our findings with regard to stroke survivors and short sleep raises the question of whether there was an adequate sample size of stroke survivors in our multivariate analyses to detect an association with mortality across all sleep duration groups, especially with the modest effect sizes expected. The sample size of 1,488 stroke survivors in our study is about half the sample size included in the single-site Hwan Kim et al study, which included 2,846 individuals.^3^ That study noted a U-shaped independent association between sleep duration and mortality. Lack of statistical power in our study is unlikely to explain our disparate findings, given our HRs are close to 1. Our study’s sample size is also significantly more than another study in a MI population, which also reported a U-shaped association between sleep duration and mortality, but only included 407 patients.^2^ Additionally, the study populations of prior studies also reporting null effects were not incongruously small.^23^ Thus, it seems unlikely that our null findings with regard to short sleep duration were due to lack of power.

The divergence in our study’s findings could have also been partially explained by the fact that the reference group (or normal duration of sleep) in our study was defined as 7 to 8 hours. The few prior studies focused on an MI population and exploring sleep duration and mortality defined normal sleep time as 6 to 8 hours.^2,3^ In prospective epidemiologic studies in the general population, various definitions of normal ranges are used. In some studies, a sleep time of 6 hours is considered too short,^25^ whereas 9 hours is in some cases considered normal.^26,27^ We thus conducted sensitivity analyses with alternate cut-off points of short sleep defined as less than 6 hours, normal sleep duration 6 – 9 hours, and greater than 9 hours of sleep comprising long duration. There were similarly no significant associations between short sleep duration and mortality among stroke survivors (or with long sleep duration and cardiovascular mortality). It is therefore less plausible that these differences in definitions could help explain our dissimilar findings.

The putative biologic mechanisms underlying an association between the extremes of sleep duration and mortality are not well understood. Previously conducted prospective epidemiologic studies have identified associations between short sleep and diabetes,^28^ obesity,^29^ and hypertension, and these may provide a credible connection between short sleep and mortality. However, the basis of the association between long sleep duration and increased mortality found in our study, as well as in epidemiologic studies,^19,30,31^ remains somewhat of a mystery, as there is little evidence to suggest that sleeping for longer than 8 hours has adverse health effects. No published basic studies or epidemiologic studies have demonstrated a possible mechanism identifying long sleep as a cause of mortality.^32^

It should be noted that even among the prospective studies finding significant associations in the general population, effect sizes have been modest. The largest study on sleep duration and mortality – the Cancer Prevention Study II^33^ – reported significant effect sizes ranging from adjusted HR = 1.07 for women who reported a sleep duration of 5 hours per night to a maximum adjusted HR = 1.41 for women who reported a sleep duration of 10 or more hours per night. The Cancer Prevention II^33^ study controlled for many demographic variables and co-morbidities, and the authors suggested that much of the mortality risk noted in association with short sleep could be explained by comorbidities.

While several prior large epidemiologic studies have investigated associations between short and long sleep duration and mortality, the strength of any associations in several of these papers is consistently several magnitudes higher for long sleep duration,^19,34,35^ than for short sleep duration, despite the lack of a clear biologic underpinning for long sleep duration. In our study, long sleep duration, but not short sleep duration, similar to some previous prospective longitudinal studies in the general population,^35^ was independently associated with higher all-cause mortality in the adjusted models. In our sensitivity analysis, short sleep duration’s initial significant association with lower cardiovascular mortality was attenuated after adjusting for demographic variables, and once comorbidities were added to the model, the association was no longer significant. Studies in the literature on this topic vary in the number of demographic variables and comorbidities that they controlled for, and so it is possible that the divergent results among studies could be partially explained by insufficient adjustment of residual confounding by incompletely measured or unmeasured variables.

In our study, we found that stroke survivors with long sleep duration (versus normal) were more likely to be older, depressed and overweight, and those with short sleep duration (versus normal) were more likely to be younger, depressed, non-White, and obese. These findings are consistent with prior large, longitudinal epidemiologic studies in the general population that have noted associations between short sleep duration and obesity,^36–38^ short sleep time among minoritized groups in comparison to white individuals,^25,39^ low socioeconomic status and long sleep,^40^ and older participants and long sleep duration.^3,25^ Our study did not detect a difference in mortality between sexes in stroke survivors, and differences among sexes in mortality outcomes related to sleep duration have been reported inconsistently in prior studies in the general population.^34^

Important strengths of this study are its large sample size, as well as adjustment in analyses for several demographic and clinical variables, however, a number of limitations must be considered. First, NHANES relies on self-reported sleep duration.

Self-reports about sleep are subject to error from imprecision based on poor recall or inadequate reporting.^41^ For example, in a prospective study (Chicago Site of Coronary Artery Risk Development in Young Adults), Lauderdale et al found that actigraph-measured sleep duration was shorter than participant self-reported sleep duration (6.06 hours versus 6.83 hours).^42^ Similar findings were reported by investigators of the Sleep Heart Health Study, which compared self-reported sleep duration to sleep duration measured using polysomnography in healthy adults.^43^ However, it is not usually feasible to obtain objective measures of sleep in large prospective, population studies. Second, NHANES surveys lack repeated measures of sleep duration; therefore, we are relying on one self-reported sleep measure, without knowing if it changed over time. Changes in sleep patterns over the follow-up period could have strengthened an association between reported sleep duration and mortality. Also, the survey does not allow for differentiation between time asleep from time in bed, or to estimate number and duration of naps. Participants with sleep disorders could be more likely to report short sleep durations due to poor sleep quality, or, conversely could be more likely to report long sleep durations due to increased time in bed to compensate for poor sleep quality. Next, NHANES survey data is likely not representative of all stroke survivors since persons with severe limitations after strokes (and those institutionalized) would likely not be included. Another limitation is the inability to classify type of stroke or stroke location in NHANES. Finally, stroke history in NHANES is based on participant report. Stroke self-report has been found to have a positive predictive value of 79%, sensitivity of 80%, and specificity of 99%.^44^

In conclusion, this study finds an independent association between long sleep duration and all-cause mortality among stroke survivors in a large nationally representative United States sample. Future longitudinal studies are needed to replicate these findings and to explore the causal links between stroke by type and location, and duration of sleep, to improve outcomes after stroke.

## Data Availability

The authors confirm that the data supporting the findings of this study are available within the article. Raw data that support the findings of this study are available from the corresponding author, upon reasonable request.

