## Supplementary figures and images for "Sleep Duration and Long-Term Mortality After Stroke: A Nationwide Analysis"

### Figure 1

**Figure 1.** Flow Chart for NHANES Stroke Survivors

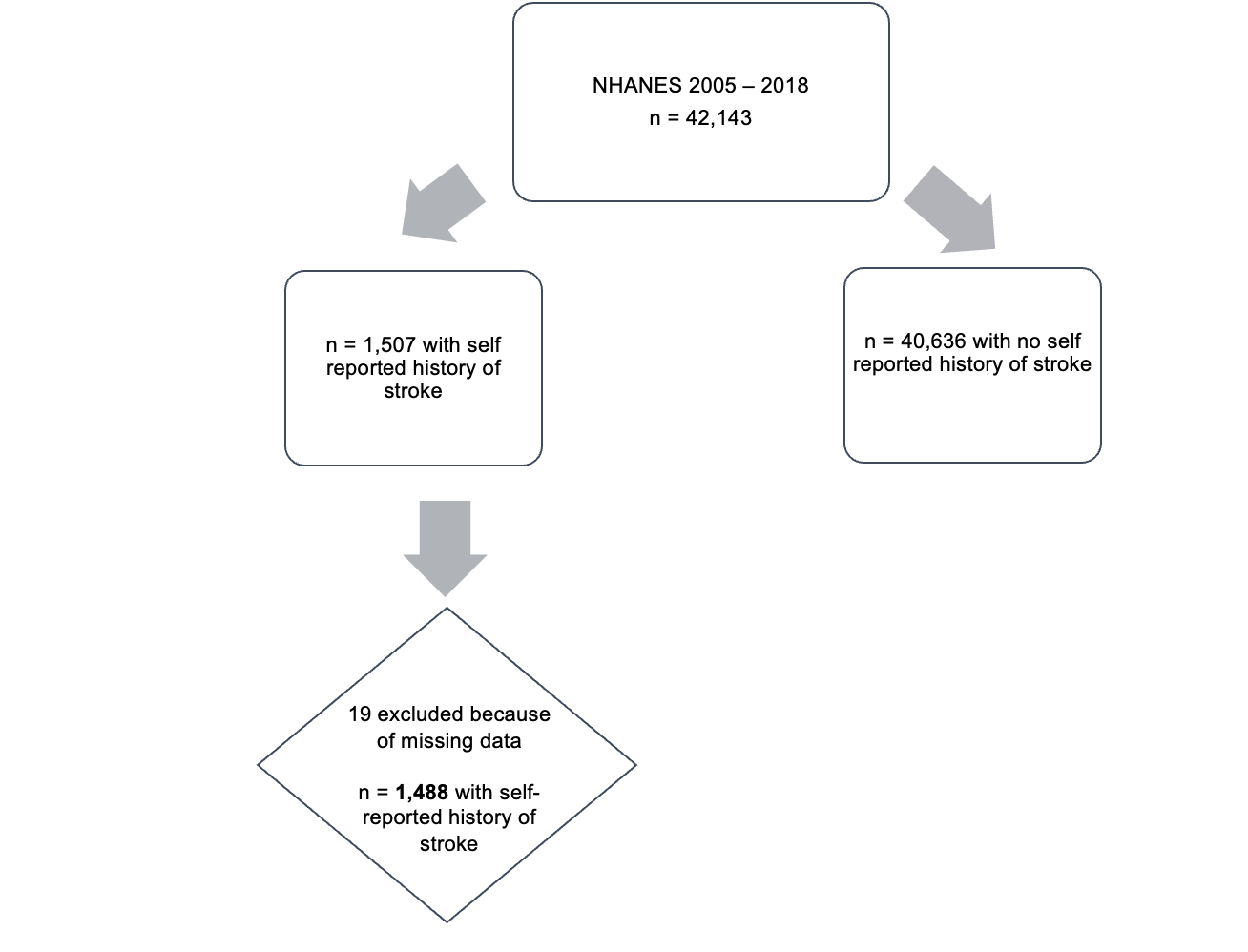
